# Prevalence of undiagnosed hypertension and prehypertension among bankers of Bangladesh: a cross-sectional study

**DOI:** 10.1101/2023.01.09.23284329

**Authors:** Sira Jam Munira, Mohammad Jahid Hasan, Md Abdur Rafi, Shafia Shaheen, Md. Iqbal Kabir

## Abstract

**Objective:** Bankers lead a sedentary and highly stressful life that often leads to developing noncommunicable diseases (NCDs) such as hypertension, diabetes, mental disorders, etc. The study aims to assess the prevalence of undiagnosed hypertension and prehypertension among bankers in Bangladesh.

**Methods:** Data from 365 bankers from five public and private banks in Bangladesh were collected using a pretested semistructured questionnaire. Prehypertension was defined as having systolic blood pressure of 120-139 mmHg and diastolic blood pressure of 80-89 mmHg. Multivariate logistic regression models were created to investigate the factors associated with them.

**Results:** The prevalence of undiagnosed hypertension and prehypertension were 22.5% and 55.3%, respectively. Most of the bankers were males and 35-44 years of age. The risk of hypertension and prehypertension was significantly higher among males (OR, 16.59; OR, 6.42), longer duration of services (F, 3.56), prolonged working hours (OR, 3.8; OR, 3.09), smoking (OR, 6.18; OR, 3.43), overweight (OR, 6.81; OR, 2.41) and obese (OR, 8.94; OR, 3.36) bankers, respectively. After controlling for all confounders, the predictors of hypertension were males (aOR, 12.8; CI, 2.73-60.02), current smokers (aOR,2.87; CI, 1.03-8), overweight (aOR,5.11; CI, 1.46-17.93), and obesity (aOR, 9.59; CI, 2.41-38.22). For prehypertension, males (aOR, 9.72; CI, 3.06-30.87) and obesity (aOR, 3.95; CI, 1.52-10.25) were found as predictors.

**Conclusion:** More than three fourth of bankers in Bangladesh have either undiagnosed hypertension or prehypertension associated with several contributing factors to occur. A large-scale study is recommended to understand the clear picture of the overall NCD risk factors burden among bankers in Bangladesh.

**WHAT IS ALREADY KNOWN ON THIS TOPIC:** The prevalence of hypertension and prehypertension are already higher in Bangladesh. Sedentary lifestyles and work stress make bankers a vulnerable community to developing these NCD risk factors.

**WHAT THIS STUDY ADDS:**

- Bankers have a higher prevalence of undiagnosed hypertension and prehypertension in Bangladesh.
- Several factors gender, long duration of service, prolonged working hours, and physical inactivity leading to overweight or obesity were found to be significant predictors of developing them.

**HOW THIS STUDY MIGHT AFFECT RESEARCH, PRACTICE OR POLICY:** This research suggests that it is now a national demand to conduct large scale surveys to assess the NCD risk factors burden among high risk occupational groups like bankers.

## INTRODUCTION

Raised blood pressure is one of the leading causes of global disease burden, contributing to almost 7% of global disability-adjusted life years (DALYs), especially in middle- and lower-income countries ^1^. The World Health Organization declared hypertension a ‘Silent Killer-Global Public Health Crisis’ ^2^. Hypertension is attributable to more than 9.4 million deaths annually due to cardiovascular diseases, which is the leading cause of death worldwide^2^. Prehypertension is considered a precursor of clinical hypertension which contributed to half of the disease burden ^3^. Both enhance the risk of cardiovascular disease-related mortality. The most recent estimates suggest that the prevalence of prehypertension in adults ranges from 17.9 to 22.2, with significant variation in rural and urban residences ^4^. However, the actual prevalence of hypertension and prehypertension among Bangladeshi bankers is not known and genderwise or occupationwise prevalence is not ready at hand.

Over the years, several risk factors for prehypertension and hypertension have been identified. These factors can be classified as nonmodifiable factors, such as age, sex, family history, etc., and modifiable factors, such as physical activity, tobacco use, alcohol and caffeinated drink intake, table salt, and saturated fat intake, nutritional status, etc. ^3,5^. Among them, obesity with hypercholesteremia is notably linked with prehypertension ^6,7^.

Raised blood pressure is one of the most common self-reported occupational health problems and is found to be significantly higher among bankers ^8^. The major contributors were increased workload, job insecurity, and extra work hours. Additionally, a sedentary lifestyle, fatty, and junk food-rich diet during office work, smoking, alcohol intake, physical inactivity, prolonged sitting posture, and a highly stressful working environment result in prehypertension and hypertension. Several studies in different geographical locations also support the epidemiological link between a sedentary lifestyle and the risk of prehypertension and conversion to hypertension ^9^. In India, the reported prevalence of prehypertension among bankers ranged from 23% to 42%, and hypertension was 31% to 50% ^10–12^. Data from low socioeconomic countries (Nigeria) ^13^ and higher socioeconomic countries (Brazzaville and Belgium) ^14,15^ also showed a similar rate of prevalence of prehypertension and hypertension among bankers.

Despite being a high risk group, evidence on the prevalence of hypertension ad prehypertension among bankers in Bangladesh is scanty. In this context, estimating the prevalence of undiagnosed hypertension and prehypertension among bankers in Bangladesh was the primary objective of the study.

## METHODS

### Study design, sampling technique, and eligibility criteria

This cross-sectional study was conducted in two public and three private banks in the capital of Bangladesh from 1^st^ January to 31^st^ December 2018. The sample size was estimated using the following formula: **n = z**^**2**^**p(1-p)/d**^**2**^, where z is 1.96 at a 95% confidence level, p is the prevalence of prehypertension among bankers, which was estimated as 34.5% from a previous study ^12^, and d (margin of error) was 5%. Considering these statistics, the calculated sample size was 365 for the present study. Simple random sampling technique was used for recruiting the participants.

The inclusion criteria were bankers who worked for ≥6 months, were designated as an officer or above the rank, and were willing to participate in the study. Diagnosed cases of hypertension and/or previous history of coronary heart disease, congestive heart failure, myocardial infarction, history of stroke, and cancer refrained from inclusion. In addition, women with current pregnancy were also excluded from the study.

### Data collection

Data were collected using a pretested semistructured questionnaire. The International Physical Activity Questionnaire (IPAQ) ^16,17^ and the Effort Reward Imbalance (ERI) ^18^ scale were used to assess physical activity and stress, respectively. The questionnaire was developed in English and then translated into Bengali. Following the pretesting on 20 subjects, necessary changes were made accordingly. The lead author collected the data through a face-to-face interview. Each interview lasted approximately thirty minutes. Informed written consent was obtained before the enrollment of the participants.

### Measurement of variables

#### Blood pressure

Blood pressure was measured using an aneroid sphygmomanometer and Littman classic III stethoscope. Before measuring blood pressure, every banker was asked to sit in a relaxed position for at least 5 minutes in a chair, feet on the floor, back supported, legs uncrossed, and arms supported on the table at heart level. The cuff was placed on encircling 80% or more of the upper left arm and ensuring that no tight clothing constricted the arm. Then, both blood pressures were recorded. Another blood pressure reading was taken after 5 minutes on the right arm. If there was a 10 mm Hg or more difference between the two readings, a 3^rd^ reading was taken. It was also ensured that the respondent did not drink tea or coffee, smoke cigarettes, or do any physical exercise thirty minutes before measuring blood pressure. The average of two measurements was recorded as systolic blood pressure (SBP) and diastolic blood pressure (DBP). Prehypertension was defined as an SBP of 120-139 mmHg and/or a DBP of 80-89 mmHg. Hypertension was defined as systolic BP≥140 mmHg and/or diastolic BP ≥90 mmHg ^19^.

#### Anthropometric measurements

The investigator measured height using a wall-mounted stadiometer and weight using a digital weighing machine. The waist and hip circumferences were measured with a measuring tape. For BMI (Body mass index), the cut-off points were >22.9 kg/m^2^ for overweight and ≥ 27.5 for obesity. For measuring central obesity, the cut-off points for waist circumference (WC) were ≥90 cm (men) and ≥80 cm (women), and for waist-hip ratio (WHR) were ≥0.90 cm (men) and ≥0.85 cm (women). Health risks were determined according to WHR, and the cut-off points were ≥1 cm (men) and ≥0.86 cm (women).

#### Physical activity

Physical activity was measured by the short version of IPAQ and classified by computing MET (metabolic equivalent of task) minutes. Three categories were inactive (no activity or **s**ome activity but not enough to be minimally active or HEPA active), minimally active (5 or more days of any combination of walking, moderate or vigorous intensity activities achieving a minimum of at least 600 MET minutes/week) and HEPA (health-enhancing physical activity) active (vigorous-intensity activity on at least three days and accumulating at least 1500 MET minutes/week **or** seven or more days of any combination of walking, moderate or vigorous intensity activities achieving a minimum of at least 3000 MET minutes/week) ^16^.

#### Work stress

The ERI scale was used for measuring stress in original English and classified by computing the effort-reward (ER) ratio. An ER ratio ≤1 was defined as low stress, and >1 was defined as high stress ^20^.

#### Statistical analysis

Statistical analyses were performed using SPSS 24 software. Frequency and percentage were used to express categorical variables, whereas mean and standard deviation were used to describe continuous, discrete variables. A chi-square test and one-way ANOVA were performed wherever applicable. A multinomial logit model (an arbitrary p value <0.05 to include variables) was created to identify risk factors for prehypertension and hypertension among bankers after controlling for all confounders. Odds ratio (OR) and adjusted Odds ratio (aOR) were calculated to assess the risk factors. Statistical significance was set at p<0.05 in a 95% confidence interval (CI).

## RESULTS

### Characteristics of bankers

A mixture of employees from private (51.5%) and public (48.5%) banks was included in the present study, wherein the majority were male (79%). The mean age was 37 (SD, 7.5) years. The average duration of service in banking was 11.2 (SD 7.8) years. Almost half of the bankers worked excess hours (47%), had inadequate sleep (65%), and were physically inactive (47%). Among lifestyle-related factors, 29% were tobacco users, 12% were alcoholics, and 22% consumed extra table salt with foods. Furthermore, a large number of bankers were found overweight (57%) and obese (28%) by BMI. About 71% and 88.5% of bankers were found centrally obese considering WC and WHR, respectively. **(Table 1)**.

**Table 1:**
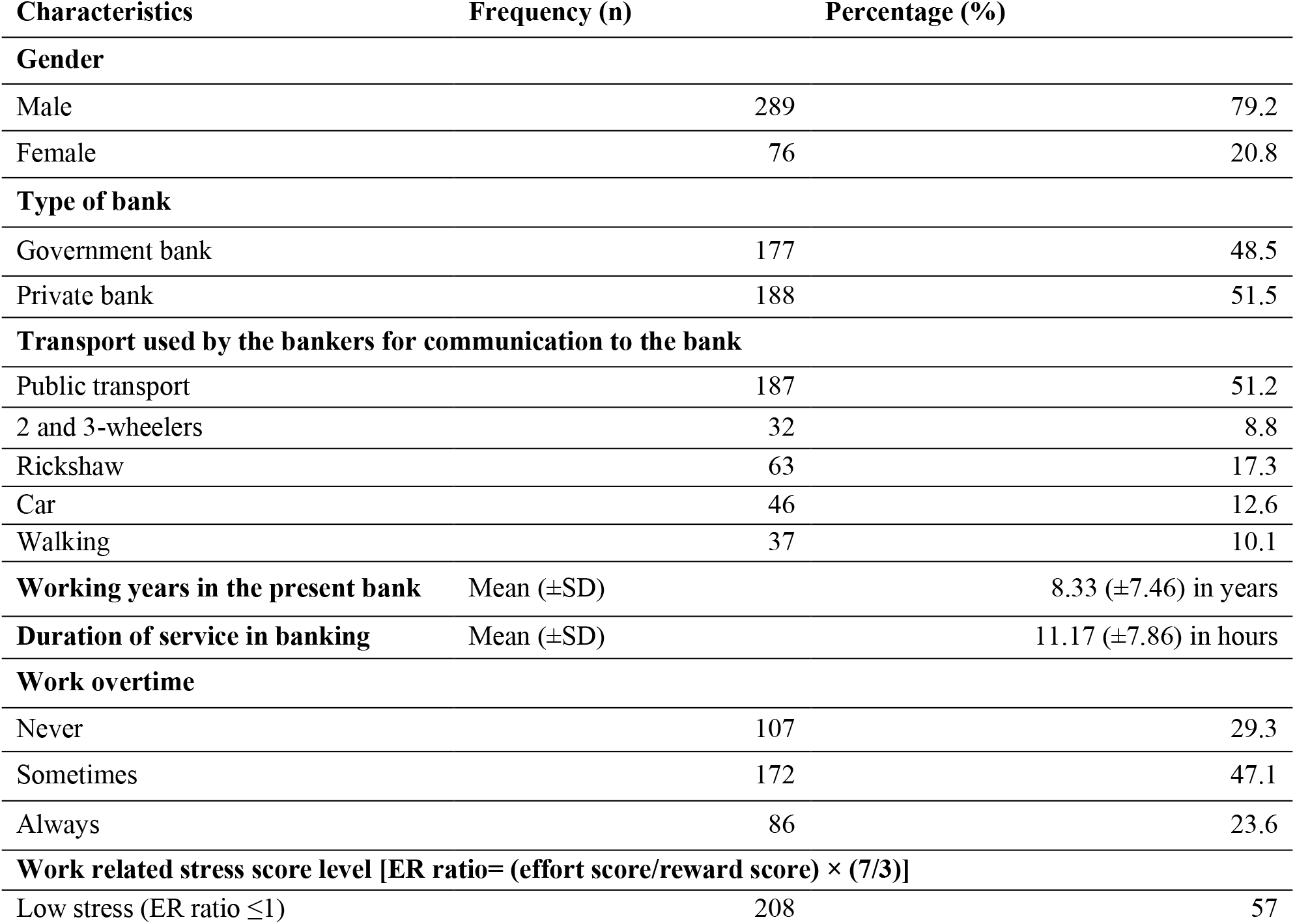
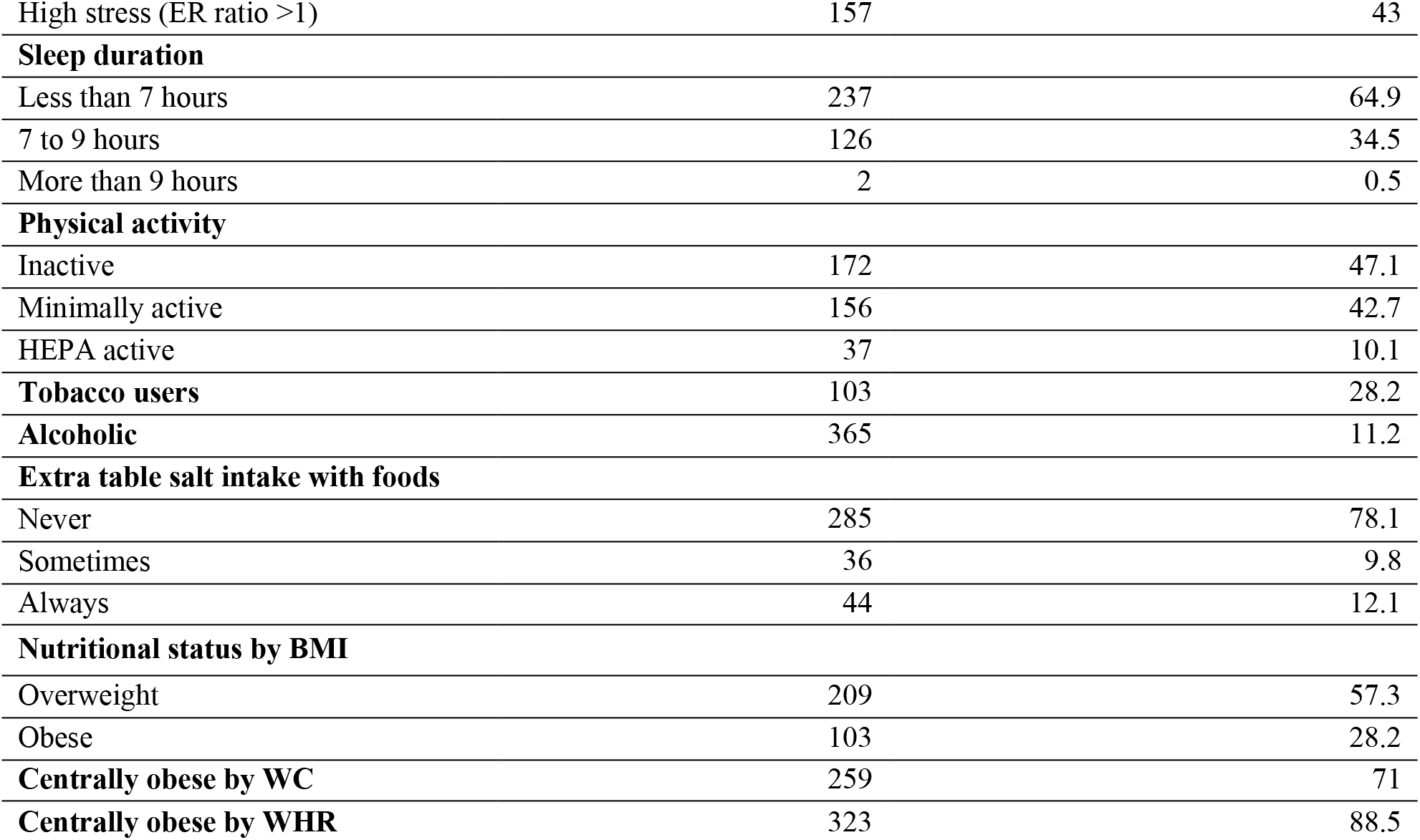
Characteristics of the bankers according to sociodemographic, banking work, and lifestyle-related factors (N= 365)

| Characteristics | Frequency (n) | Percentage (%) |
| --- | --- | --- |
| **Gender** |  |  |
| Male | 289 | 79.2 |
| Female | 76 | 20.8 |
| **Type of bank** |  |  |
| Government bank | 177 | 48.5 |
| Private bank | 188 | 51.5 |
| **Transport used by the bankers for communication to the bank** |  |  |
| Public transport | 187 | 51.2 |
| 2 and 3-wheelers | 32 | 8.8 |
| Rickshaw | 63 | 17.3 |
| Car | 46 | 12.6 |
| Walking | 37 | 10.1 |
| **Working years in the present bank** | Mean (±SD) | 8.33 (±7.46) in years |
| **Duration of service in banking** | Mean (±SD) | 11.17 (±7.86) in hours |
| **Work overtime** |  |  |
| Never | 107 | 29.3 |
| Sometimes | 172 | 47.1 |
| Always | 86 | 23.6 |
| **Work related stress score level [ER ratio= (effort score/reward score) × (7/3)]** |  |  |
| Low stress (ER ratio ≤1) | 208 | 57 |
| High stress (ER ratio >1) | 157 | 43 |
| **Sleep duration** |  |  |
| Less than 7 hours | 237 | 64.9 |
| 7 to 9 hours | 126 | 34.5 |
| More than 9 hours | 2 | 0.5 |
| **Physical activity** |  |  |
| Inactive | 172 | 47.1 |
| Minimally active | 156 | 42.7 |
| HEPA active | 37 | 10.1 |
| **Tobacco users** | 103 | 28.2 |
| **Alcoholic** | 365 | 11.2 |
| **Extra table salt intake with foods** |  |  |
| Never | 285 | 78.1 |
| Sometimes | 36 | 9.8 |
| Always | 44 | 12.1 |
| **Nutritional status by BMI** |  |  |
| Overweight | 209 | 57.3 |
| Obese | 103 | 28.2 |
| **Centrally obese by WC** | 259 | 71 |
| **Centrally obese by WHR** | 323 | 88.5 |

### Prevalence of undiagnosed hypertension and prehypertension

Undiagnosed hypertension and prehypertension were detected in 22.5% and 55.3% of the bank employees respectively. The prevalence of systolic hypertension and prehypertension 5% and was 45%, respectively while the prevalence of diastolic hypertension and prehypertension was 22% and 55%, respectively **(Figure 1)**.

**Figure 1:**
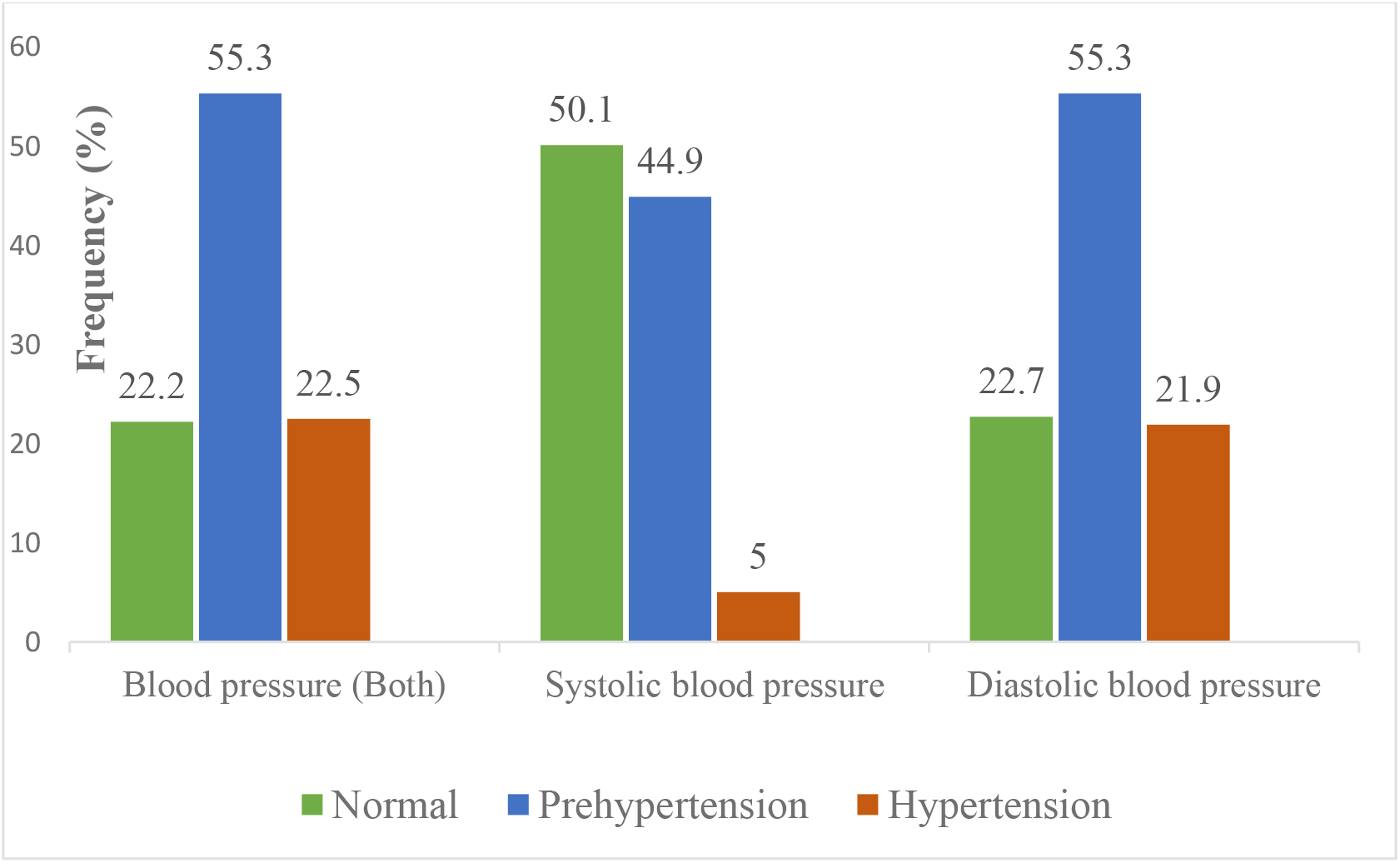
Distribution of the bankers by blood pressure level (N=365)

### Factors associated with undiagnosed hypertension and prehypertension

Approximately, 26.6% of male bankers and 6.6% of female bankers were hypertensive, and 59.9% of male bankers and 38.2% of female bankers were prehypertensive (p<0.001). Among the banking work-related factors, work overtime (p<0.01) and longer duration of service in banking (p<0.05) were associated with both undiagnosed hypertension and prehypertension. Approximately 59.2% obese, 55% overweight (p<0.01) by BMI, and 55.7% centrally obese (p<0.05) by WHR were prehypertensive. For undiagnosed hypertension 25.2% were obese, 24.9% were overweight (p<0.01) by BMI, and 25.1% & 24.1% were centrally obese (p<0.05) by WC and WHR, respectively. **(Table 2)**.

**Table 2:** Association of sociodemographic, banking work-related factors, and anthropometric measurements with blood pressure level (N=365)

| Characteristics | Categories | Normotension<br>n (%) | Prehypertension<br>n (%) | Hypertension<br>n (%) | P |
| --- | --- | --- | --- | --- | --- |
| Age | 25-34 years | 45 (28.7) | 85 (54.1) | 27 (17.2) | 0.093 |
|  | 35-44 years | 27 (18.4) | 82 (55.8) | 38 (25.9) |  |
|  | 45- 54 years | 6 (13.3) | 28 (62.2) | 11 (24.4) |  |
|  | ≥ 55years | 3 (18.8) | 7 (43.8) | 6 (24.4) |  |
| Sex | Male | 39 (13.5) | 173 (59.9) | 77 (26.6) | <0.001 |
|  | Female | 42 (55.3) | 29 (38.2) | 5 (6.6) |  |
| Educational status | Graduate | 9 (40.9) | 10 (45.5) | 3 (13.6) | <0.05 |
|  | Postgraduate | 46 (18.3) | 150 (59.5) | 56 (22.2) |  |
|  | Professional (Engineering) | 26 (28.6) | 42 (46.2) | 23 (25.3) |  |
| Type of bank | Govt. bank | 47 (26.6) | 88 (49.7) | 42 (23.7) | 0.076 |
|  | Private bank | 34 (18.1) | 114 (60.6) | 40 (21.3) |  |
| Transport used for communication to the bank | Public Transport | 44 (23.5) | 98 (52.4) | 45 (24.1) | 0.268 |
|  | 2 and 3-wheelers | 17 (17.9) | 57 (60) | 21 (22.1) |  |
|  | Car | 15 (32.6) | 25 (54.3) | 6 (13) |  |
|  | Walking | 5 (13.5) | 22 (59.5) | 10 (27) |  |
| Work overtime | Never | 32 (29.9) | 59 (55.1) | 16 (15) | <0.01 |
|  | Sometimes | 39 (22.7) | 86 (50) | 47 (27.3) |  |
|  | Always | 10 (11.6) | 57 (66.3) | 19 (22.1) |  |
| Work-related stress | Low stress | 48 (23.1) | 114 (54.8) | 46 (22.1) | 0.895 |
|  | High stress | 33 (21)) | 88 (56.1) | 36 (22.9) |  |
| Duration of service [Mean (±SD)] |  | 7.03 (±7.95) | 8.97 (±7.77) | 9.42 (±7.93) | <0.05 |
| Daily working hours in sitting posture [Mean (±SD)] |  | 6.71 (±1.61) | 6.79 (±1.68) | 6.73 (±1.61) | 0.913 |
| Nutritional status by BMI | Underweight | 1 (50) | 1 (50) | 0 (0) | **<0.01** |
|  | Normal | 22 (43.1) | 25 (49) | 4 (7.8) |  |
|  | Overweight | 42 (20.1) | 115 (55) | 52 (24.9) |  |
|  | Obese | 16 (15.5) | 61 (59.2) | 26 (25.2) |  |
| Central obesity (WC) | Not obese | 26 (24.5) | 63 (59.4) | 17 (16) | 0.168 |
|  | Obese | 55 (21.2) | 139 (53.7) | 65 (25.1) |  |
| Central obesity (WHR) | Not obese | 16 (38.1) | 22 (52.4) | 4 (9.5) | **<0.05** |
|  | Obese | 65 (20.1) | 180 (55.7) | 78 (24.1) |  |

Among lifestyle-related factors, being physically inactive and minimally active (p<0.01), tobacco users (p<0.001), current smokers (p<001), weekly exposure to secondhand smoking (p<0.05) and always consuming extra table salt with cooked foods (p<0.05) were associated with undiagnosed hypertension and prehypertension. Although a positive family history of hypertension, alcoholic, regular intake of caffeinated drinks, salty and oily fast foods were not associated with them. **(Table 3)**.

**Table 3:**
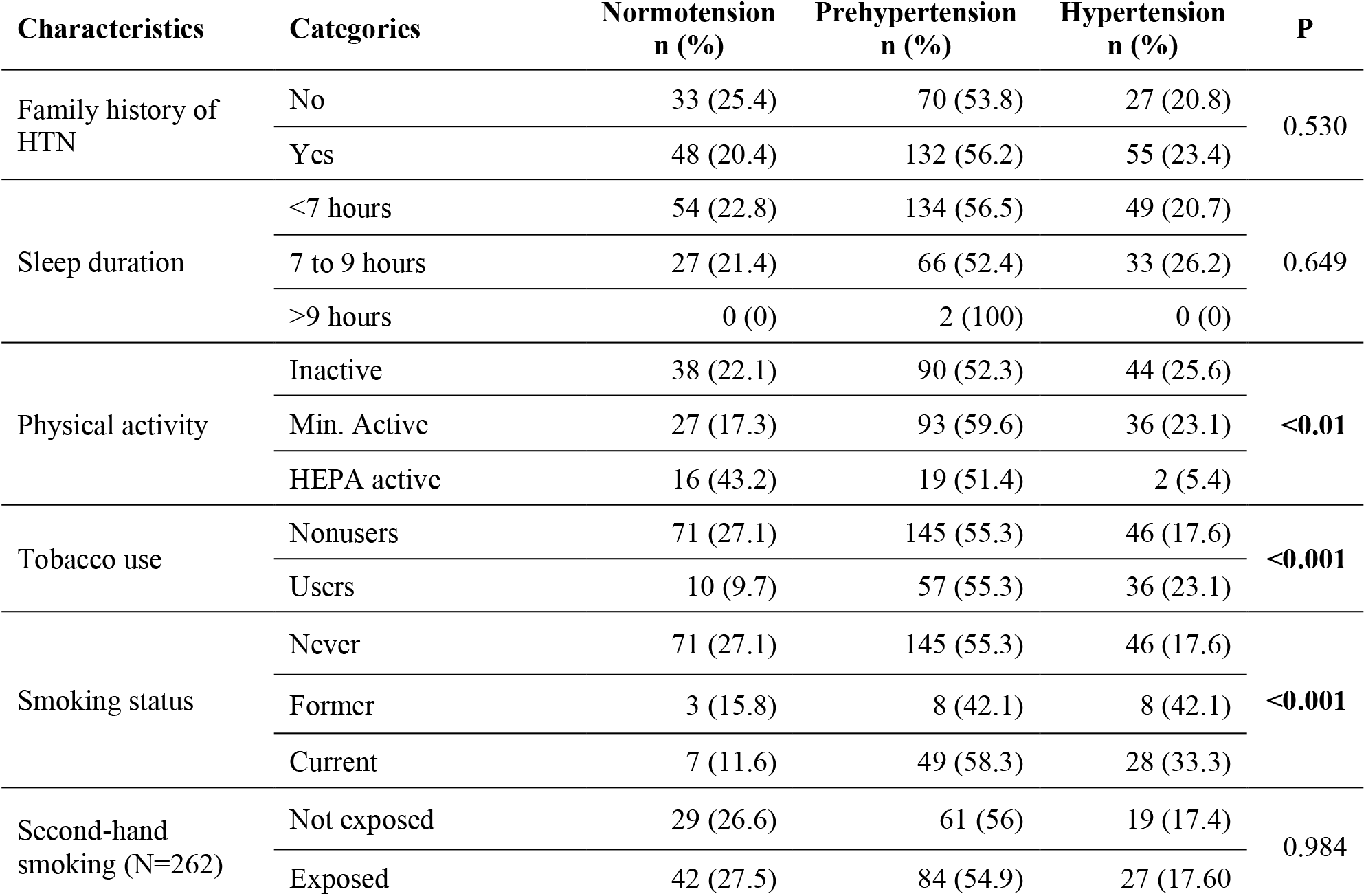
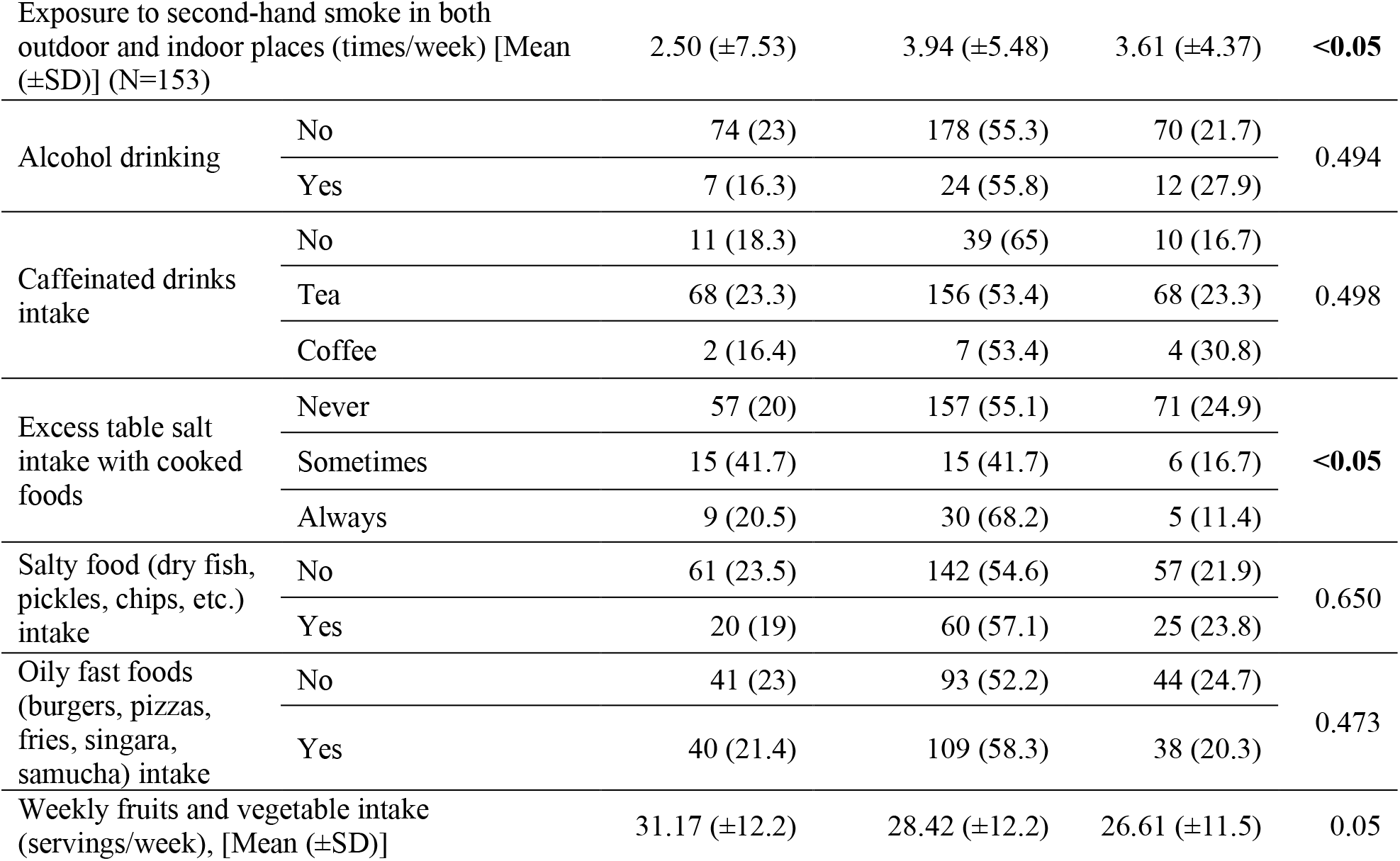
Association of lifestyle-related factors with blood pressure level (N=365)

A multivariable logistic regression model was created to predict the factors of undiagnosed hypertension and prehypertension including the variables found to be significant in the Chi square test and one-way ANOVA test. After running the multicollinearity test, tobacco product users and exposure to second-hand smoke in indoor and outdoor places were excluded from the model. We found that males (12.796 times more than females), overweight (5.11 times more than normal weight), obese (9.59 times more than normal weight), and current smokers (2.87 times more than never smokers) were significantly associated with undiagnosed hypertension among bankers. Additionally, males (9.723 times more than females) and obesity (3.947 times more than normal weight) were significantly associated with prehypertension among bankers after controlling for confounders. **(Table 4)**.

**Table 4:** Comparison of factors for prehypertension and hypertension (both vs. normal BP) groups after controlling for confounders (N=365)

| Characteristics | Categories | Prehypertension vs. Normotension |  | Hypertension vs. Normotension |  |
| --- | --- | --- | --- | --- | --- |
|  |  | aOR (95% CI) | P | aOR (95% CI) | P |
| Duration of service |  | 1.02 (0.98-1.07) | 0.371 | 1.03 (0.98- 1.09) | 0.236 |
| Sex | Female | Reference Category |  |  |  |
|  | Male | 9.72(3.06-30.87) | **<0.001** | 12.79(2.73-60.02) | **<0.001** |
| Educational status | Graduate | Reference Category |  |  |  |
|  | Postgraduate | 1.03(0.25- 4.16) | 0.971 | 2.56(0.46- 14.36) | 0.285 |
|  | Professional | 2.98(0.83- 1.69) | 0.95 | 1.21(0.19- 7.64) | 0.843 |
| Work overtime | Never | Reference Category |  |  |  |
|  | Sometimes | 0.64(0.31- 1.31) | 0.223 | 1.15(0.48- 2.79) | 0.754 |
|  | Always | 1.21(0.46- 3.16) | 0.699 | 1.24(0.39- 3.89) | 0.715 |
| Physical activity | Inactive | Reference Category |  |  |  |
|  | Min. active | 1.26(0.65- 2.45) | 0.503 | 0.97(0.45- 2.07) | 0.93 |
|  | HEPA active | 1.61(0.581- 4.45) | 0.361 | 0.47(0.08- 2.73) | 0.40 |
| Smoking status | Never | Reference Category |  |  |  |
|  | Former | 0.54(0.13- 2.38) | 0.419 | 1.63(0.36- 7.32) | 0.526 |
|  | Current | 1.64(0.63- 4.28) | 0.309 | 2.87(1.03- 8.01) | **<0.05** |
| Excess table salt use | Never | Reference Category |  |  |  |
|  | Sometimes | 0.49(0.19- 1.28) | 0.148 | 0.45(0.15- 1.63) | 0.247 |
|  | Always | 1.61 (0.63- 4.13) | 0.319 | 0.68(0.19- 2.46) | 0.553 |
| Nutritional status by BMI | Normal | Reference Category |  |  |  |
|  | Underweight | 0.65(0.02- 18.26) | 0.802 | Not valid |  |
|  | Overweight | 1.82(0.81- 4.10) | 0.150 | 5.11(1.46- 17.93) | **<0.05** |
|  | Obese | 3.95(1.52- 10.25) | **<0.01** | 9.59(2.41- 38.22) | **<0.001** |
| Central obesity by WHR | Not obese | Reference Category |  |  |  |
|  | Obese | 1.40(0.55- 3.57) | 0.481 | 2.18(0.57- 8.41) | 0.257 |

## DISCUSSION

This cross-sectional study was conducted to determine the proportion of bankers with prehypertension and hypertension and the factors associated with prehypertension The proportions of prehypertension and hypertension were 55.3% and 22.5%, respectively. The overall prevalence of prehypertension and hypertension in an affluent urban area of Dhaka was 19% and 23.7% ^21^, among the general population of Bangladesh was 43% and 20.1% ^4^. If we consider bankers of several provinces in India, the overall prevalence of prehypertension and hypertension was 34% -42% and 30% - 45% ^10,22,23^. Our findings suggested that the prevalence of prehypertension among bankers was higher compared to the general population of urban and rural areas in Bangladesh and Indian bankers. However, regarding undiagnosed hypertension, bankers and the general population of Bangladesh had almost the same level, and Indian bankers had a higher level.

We found that the proportion of undiagnosed hypertension and prehypertension was higher in males. However, no association was found with increasing age. Several studies showed a significant association between sex and age and BP levels in Bangladeshi ^4,24^, Indian ^25^, Chinese ^26^, Japanese ^27^, and Iranian ^28^ adult populations and bankers. Sex and age are the two major predictors of major hypertensive disorders ^29,30^. In our studies, we could not find a significant association with age; perhaps the absence of age uniformity among bankers is a reason for this. The proportion of undiagnosed hypertension and prehypertension was higher in bankers with long years of schooling, which corresponds with Bangladeshi ^4^ and Indian studies ^31^.

Among banking work-related factors, we found that prolonged working hours and long duration of service in banking had a significant association with both levels, which corresponded with some study findings ^9,32,33^. Several studies have shown that work stress is associated with hypertension and prehypertension ^10^. However, we could not find any relationship between mental stress, prehypertension, and hypertension. Major reasons are hiding information from the bankers’ professional point of view, minor variation in sampling, and job environment.

Among several anthropometric measurements, we found a significant association with BMI, prehypertension, and hypertension levels among bankers. These findings are similar to the finding that overweight and obesity are the strongest predictors of the higher prevalence of prehypertension and hypertension among Bangladeshi ^4^, Indian ^25^, Chinese ^26^, Iranian ^28^, Jamaican ^34^, and Japanese ^27^ adult populations. Central obesity measured by WC showed no association, and WHR was significantly associated with prehypertension and hypertension among bankers. However, some studies showed a significant association with WC, WHR, and BP levels ^34,35^.

In this study, lifestyle-related factors associated with prehypertension and hypertension were explored. Many studies suggest that family history plays a significant role in developing hypertension in adults ^27,36^. Shorter sleep duration increases BP levels ^33^, a risk factor for CVDs. This study’s findings showed no association. We found that more physical activity was inversely associated with prehypertension and hypertension, which corresponded with several studies’ findings among the general population ^25,37^ and bankers ^23^.

Tobacco product users and current smokers were at an increased risk of developing prehypertension and hypertension among bankers. These findings corresponded with several studies’ results among the general population ^25,31^ and bankers ^10,38^. The average weekly exposure to second-hand smoking (times/week) in both places (indoor and outdoor) was higher in bankers with prehypertension and hypertension, which corresponded with the remarkable acute effect of second-hand smoking on blood pressure levels.

Many studies on the adult population showed that heavy alcohol consumption ^25,27,31,35^ and increased caffeine ingestion ^39^ increased BP levels. Although the present study showed no association, it might be due to low alcohol consumption for religious reasons or our social customs.

The proportion of prehypertension was higher in 68.2% of bankers who always took extra table salt with cooked foods, which corresponded with excess table salt intake with foods increasing blood pressure levels and the risk of CVDs ^40^. Goswami and Narayan’s study on India showed that consuming high-energy food such as salty, oily junk food increased BP levels ^31^. However, this study showed no association. A study in India among bankers showed that a low intake of fruits and vegetables increased hypertension levels ^10^. However, this study showed no significant association.

A multinomial logistic regression model was created to predict factors of prehypertension and hypertension among bankers. After controlling for all confounders, we found that male gender and obesity or overweight were associated with both undiagnosed hypertension and prehypertension. Current smoking showed a significant association with hypertension only. The WHO steps survey ^4^ and Khanam’s study on rural people ^24^ of Bangladesh showed that prehypertension and hypertension were associated with male gender, older age groups, higher education, and higher BMI. Two studies in India among bankers showed that males, increasing age, increased mental stress, family history of hypertension, low physical activity, smoking, caffeinated drinks, and excess salt intake with foods increased the risk of hypertension ^10,23^. This study’s findings were inconsistent with all these factors, but only a few showed significant associations.

### Strengths and limitations

The main strengths of this study were to determine the prevalence of undiagnosed hypertension and prehypertension along with the associated factors and probably the first of its kind among bankers in Bangladesh. The weakness of this study was the nonrepresentative banking community of the whole country and the small sample size. This study was conducted in five selected banks in Dhaka, so the findings cannot be generalized to all bankers in Bangladesh. Additionally, there might be some bias, as BP was measured in a single setting. The presence of some potential risk factors was objectively measured based on memory leading to recall bias. There was also some social desirability bias due to hiding information from the bankers’ professional point of view.

## CONCLUSIONS

This study suggests that bankers are highly vulnerable professionals to hypertension and prehypertension. Almost half of them were prehypertensive, and nearly one-fourth of them were unaware of their hypertensive condition. Male gender, overweight or obesity, and smoking habits increase the risk of both hypertension and prehypertension. Adequate preventive measures are suggested to prevent the high burden of these NCDs.

## Data Availability

All data produced in the present study are available upon reasonable request to the authors.

## ACKNOWLEDGEMENTS

The authors thank the Department of Epidemiology, National Institution of Preventive and Social Medicine (NIPSOM) for their continuous support and guidance. The authors place their profound gratitude to Md. Anowar Hossain, DGM, Human Resource Department, Janata Bank Limited, and Md. Mamtaz Uddin Chowdhury, Vice President, Head of Agriculture Loan Unit, Jamuna Bank Limited. This study would not have been possible without the positive attitude, cooperation, and willingness of the bankers of selected banks.

## Contributors

SJM and MIK planned the study, data analysis, and statistics. SJM collected data, analyzed it, and wrote the first draft of the manuscript. SJM, MJH, MAR, and SS revised the manuscript and provided final edits. All authors read and approved the final manuscript.

## Funding

Not applicable.

## Ethical approval

Formal ethical clearance for the protocol of the study was obtained from the Institutional Review Board (IRB) of NIPSOM (Memo no: NIPSOM/IRB/2018/471).

## Informed written consent

Informed written consent was ensured from each participant before participation in the study. The study was confirmed by the ethical guidelines of the current Declaration of Helsinki.

## Patient consent for publication

Not applicable.

## Data availability statement

The data collection process is described in the method section of this article. All data and details from the analysis can be obtained from the corresponding author on request.

## Conflict of interests

There is no conflict of interest.

## Notes

### Competing Interest Statement

The authors have declared no competing interest.

### Funding Statement

This study did not receive any funding

### Author Declarations

Institutional Review Board of National Institute of Preventive and Social Medicine, Dhaka, Bangladesh gave ethical approval for this work.

## REFERENCES

1. Lim SS, Vos T, Flaxman AD, et al. A comparative risk assessment of burden of disease and injury attributable to 67 risk factors and risk factor clusters in 21 regions, 1990-2010: A systematic analysis for the Global Burden of Disease Study 2010. Lancet. 2012;380(9859):2224–2260. doi:10.1016/S0140-6736(12)61766-8

2. WHO. A Global Brief on Hypertension: Silent Killer, Global Public Health Crisis.; 2013. doi:10.1136/bmj.1.4815.882-a

3. Vasan RS, Larson MG, Leip EP, Kannel WB, Levy D. Assessment of frequency of progression to hypertension in non-hypertensive participants in the Framingham Heart Study: A cohort study. Lancet. 2001;358(9294):1682–1686. doi:10.1016/S0140-6736(01)06710-1

4. Rahman M, Zaman MM, Islam JY, et al. Prevalence, treatment patterns, and risk factors of hypertension and pre-hypertension among Bangladeshi adults. J Hum Hypertens. 2018;32(5):334–348. doi:10.1038/s41371-017-0018-x

5. Winegarden CR. From “prehypertension” to hypertension? Additional evidence. Ann Epidemiol. 2005;15(9):720–725. doi:10.1016/j.annepidem.2005.02.010

6. Asmathulla S, Rajagovindan D, Sathyapriya V, Pai B. Prevalence of prehypertension and its relationship to cardiovascular disease risk factors in Puducherry. Indian J Physiol Pharmacol. 2011;55(4):343–350. doi:http://dx.doi.org/10.1016/j.ijcard.2011.08.783

7. Greenlund KJ, Croft JB, Mensah GA. Prevalence of Heart Disease and Stroke Risk Factors in Persons With Prehypertension in the United States, 1999-2000. Arch Intern Med. 2004;164:1999–2000.

8. Maxcy-rosenau-last, Wallace RB. Public Health & Preventive Medicine. Fifteenth. (Wallace RB, ed.). The McGraw-Hill doi:10.1036/0071441980

9. Sit JWH, Sijian L, Wong EMY, et al. Prevalence and risk factors associated with prehypertension: Identification of foci for primary prevention of hypertension. J Cardiovasc Nurs. 2010;25(6):461–469. doi:10.1097/JCN.0b013e3181dcb551

10. Brahmankar TR, Prabhu PM. Prevalence and risk factors of hypertension among the bank employees of Western Maharashtra – a cross sectional study. Int J Community Med Public Heal. 2017;4(4):1267-277. doi:10.18203/2394-6040.ijcmph20171361

11. Sumalatha N, Sumalatha N, Dorle AS, Anjum W, Gagan S. Study of Socio-demographic Profile & Prevalence Of Hypertension among Bank Employees in Bagalkot City. Ann Community Heal. 2015;3(1):28–32. http://www.annalsofcommunityhealth.in/ojs/index.php/AoCH/article/view/151

12. Momin M, Kavishwar A, Desai V. Study of socio-demographic factors affecting prevalence of hypertension among bank employees of Surat City. Indian J Public Health. 2012;56(1):44–48. doi:10.4103/0019-557X.96970

13. Salaudeen AG, Musa OI, Babatunde OA, Atoyebi OA, Durowade KA, Omokanye LO. Knowledge and prevalence of risk factors for arterial hypertension and blood pressure pattern among bankers and traffic wardens in Ilorin, Nigeria. Afr Health Sci. 2014;14(3):593–599. doi:10.4314/ahs.v14i3.14

14. Thierry G, Benjamin L, Bertrand E, et al. Prevalence rates and cardiometabolic determinants of diabetes mellitus and pre-diabetes with projected coronary heart disease at bank site of Brazzaville. World J Cardiovasc Dis. 2014;04(02):77–86. doi:10.4236/wjcd.2014.42012

15. Shivaramakrishna HR, Wantamutte AS, Sangolli HN, Mallapur M. Risk Factors of Coronary Heart Disease among Bank Employees of Belgaum City - Cross-Sectional Study. Al Ameen J Med Sci. 2010;3(2):152–159.

16. Guidelines for data processing and analysis of the International Physical Activity Questionnaire (IPAQ)-short form. Published online 2004. http://www.ipaq.ki.se

17. Craig CL, Marshall AL, Sjöström M, et al. International physical activity questionnaire: 12-Country reliability and validity. Med Sci Sports Exerc. 2003;35(8):1381–1395. doi:10.1249/01.MSS.0000078924.61453.FB

18. Siegrist J, Li J, Montano D. Psychometric properties of the effort-reward imbalance questionnaire. Ger Duesseld Univ. Published online 2014.

19. Chobanian A V., Bakris GL, Black HR, et al. Seventh report of the Joint National Committee on Prevention, Detection, Evaluation, and Treatment of High Blood Pressure. Hypertension. 2003;42(6):1206–1252. doi:10.1161/01.HYP.0000107251.49515.c2

20. Leineweber C, Wege N, Westerlund H, Theorell T, Wahrendorf M, Siegrist J. How valid is a short measure of effort–reward imbalance at work? A replication study from Sweden. Occup Environ Med. 2010;67(8):526–531.

21. Islam SMS, Mainuddin A, Islam MS, et al. Prevalence of risk factors for hypertension: A cross-sectional study in an urban area of Bangladesh. Glob Cardiol Sci Pract. 2015;2015(4):43. doi:10.5339/gcsp.2015.43

22. Ismail I, Kulkarni A, Kamble S, Rekha R, Amruth M, Borker S. Prevalence of hypertension and its risk factors among bank employees of Sullia Taluk, Karnataka. Sahel Med J. 2013;16(4):139. doi:10.4103/1118-8561.125553

23. Ganesh KS, Sundaram ND. Prevalence and Risk Factors of Hypertension among Bank Employees in Urban Puducherry, India. Int J Occup Env Med. 2014;5(2):94–100. doi:10.4103/0972-6748.144938

24. Khanam MA, Lindeboom W, Razzaque A, Niessen L, Smith W, Milton AH. Undiagnosed and uncontrolled hypertension among the adults in rural Bangladesh: Findings froma community-based study. J Hypertens. 2015;33(12):2399–2406. doi:10.1097/HJH.0000000000000712

25. Parthaje PM, Unnikrishnan B, Thankappan KR, Thapar REKHA, Fatt QK, Oldenburg B. Prevalence and Correlates of Prehypertension among Adults in Urban South India. Asia-Pacific J Public Heal. 2016;28(9):93S–101S. doi:10.1177/1010539515616453

26. Wang R, Lu X, Hu Y, You T. Prevalence of prehypertension and associated risk factors among health check-up population in Guangzhou, China. Int J Clin Exp Med. 2015;8(9):16424–16433.

27. Ishikawa Y, Ishikawa J, Ishikawa S, et al. Prevalence and Determinants of Prehypertension in a Japanese General Population: The Jichi Medical School Cohort Study. Hypertens Res. 2008;31(7):1323–1330. doi:10.1291/hypres.31.1323

28. Rahmanian K, Shojaie M. The prevalence of pre-hypertension and its association to established cardiovascular risk factors in south of Iran. BMC Res Notes. 2012;5(1):1. doi:10.1186/1756-0500-5-386

29. Sarki AM, Nduka CU, Stranges S, Kandala NB, Uthman OA. Prevalence of hypertension in low- and middle-income countries: A systematic review and meta-analysis. Med (United States). 2015;94(50):1–16. doi:10.1097/MD.0000000000001959

30. Chowdhury MZI, Rahman M, Akter T, et al. Hypertension prevalence and its trend in Bangladesh: evidence from a systematic review and meta-analysis. Clin Hypertens. 2020;26(1). doi:10.1186/s40885-020-00143-1

31. Goswami T, Narayan B. Behavioural risk factors distribution of cardiovascular diseases and its association with normotension, prehypertension and hypertension amongst tea garden population in Dibrugarh district of Assam. Clin Epidemiol Glob Heal. 2015;82:6. doi:10.1016/j.cegh.2014.10.004

32. Yoo DH, Kang MY, Paek D, Min B, Cho S Il. Effect of Long Working Hours on Self-reported Hypertension among Middle-aged and Older Wage Workers. Ann Occup Environ Med. 2014;26(1):1–10. doi:10.1186/s40557-014-0025-0

33. Cheng Y, Du CL, Hwang JJ, Chen IS, Chen MF, Su TC. Working hours, sleep duration and the risk of acute coronary heart disease: A case-control study of middle-aged men in Taiwan. Int J Cardiol. 2014;171(3):419–422. doi:10.1016/j.ijcard.2013.12.035

34. Ferguson TS, Younger NOM, Tulloch-Reid MK, et al. Prevalence of prehypertension and its relationship to risk factors for cardiovascular disease in Jamaica: Analysis from a cross-sectional survey. BMC Cardiovasc Disord. 2008;8:1–9. doi:10.1186/1471-2261-8-20

35. Liu B, Li W, Hu B, et al. Prevalence and Determinants of Prehypertension in a Chinese Population of 34-45 Years Old Heart Rate Variability and Blood Pressure Rhythm in Hypertensive Patients with Obstructive Sleep Apnea hypopnea Syndrome. 2010;23(7):2010. doi:10.1038/ajh.2010.84

36. Gyamfi D, Obirikorang C, Acheampong E, et al. Prevalence of pre-hypertension and hypertension and its related risk factors among undergraduate students in a Tertiary institution, Ghana. Alexandria J Med. Published online 2018:0–5. doi:10.1016/j.ajme.2018.02.002

37. Borjesson M, Onerup A, Lundqvist S, Dahlof B. Physical activity and exercise lower blood pressure in individuals with hypertension: Narrative review of 27 RCTs. Br J Sports Med. 2016;50(6):356–361. doi:10.1136/bjsports-2015-095786

38. Momin. M, Vikas D AK. A Study On Effect Of Life Style Risk Factors On Prevalence Of Hypertension Among White Collar Job People Of Surat - ISPUB. Internet J Occup Heal. 2011;1(2):1–9. doi:10.5580/10bc

39. Soares RN, Schneider A, Valle SC, Schenkel PC. Regular Physical Activity Increases the Systolic Blood Pressure Response to Acute Caffeine Ingestion in Nonhabitual Caffeine Consumers. J Caffeine Res. 2016;00(00):1–6. doi:10.1089/jcr.2016.0017

40. He FJ, Jenner KH, Macgregor GA. WASH — World Action on Salt and Health. Kidney Int. 2010;78(8):745–753. doi:10.1038/ki.2010.280

